# Normalization of Drug and Therapeutic Concepts with TheraPy

**DOI:** 10.1101/2023.07.27.23293245

**Authors:** Matthew Cannon, James Stevenson, Kori Kuzma, Susanna Kiwala, Jeremy L Warner, Obi L Griffith, Malachi Griffith, Alex H Wagner

## Abstract

Working with therapeutic terminology in the field of medicine can be challenging due to both the number of ways terms can be addressed and the ambiguity associated with different naming strategies. A therapeutic concept can be identified across many facets from ontologies and vocabularies of varying focus, including natural product names, chemical structures, development codes, generic names, brand names, product formulations, or treatment regimens. This diversity of nomenclature makes therapeutic terminology difficult to manage and harmonize. As the number and complexity of available therapeutic ontologies continues to increase, the need for harmonized cross-resource mappings is becoming increasingly apparent. Harmonized concept mappings will enable the linking together of like-concepts despite source-dependent differences in data structure or semantic representation. To support these mappings, we introduce TheraPy, a Python package and web API that constructs stable, searchable merged concepts for drugs and therapeutic terminologies using publicly available resources and thesauri. By using a directed graph approach, TheraPy can capture commonly used aliases, trade names, annotations, and associations for any given therapeutic and combine them under a single merged concept record. Using this approach, we found that TheraPy tends to normalize therapeutic concepts to their underlying active ingredients (excluding non-drug therapeutics, e.g. radiation therapy, biologics), and unifies all available descriptors regardless of ontological origin. In this report, we highlight the creation of 16,069 unique merged therapeutic concepts from 9 distinct sources using TheraPy. Further, we analyze rates of normalization for therapeutic terms taken from publicly available vocabularies.

## INTRODUCTION

Referencing drugs or other therapeutic terms is unavoidable in the field of medicine. Clinicians and other medical professionals are frequently expected to synthesize knowledge of drug mechanisms, effectiveness, and other metrics to design the best possible treatment regimens with their patients. Databases and other resources exist that allow medical professionals to collect information about a therapeutic of interest and synthesize this data with the literature. This process can be hampered, however, due to the difficulties in knowing exactly how therapies are being defined in each resource. Working with therapeutic terminologies can be challenging due to the number of ways some terms can be described and the ambiguity associated with different naming strategies.^1^ This problem is exacerbated in clinical genomics, where ambiguity (or a lack of standardization) can confound treatment decision making. Consider imatinib, a tyrosine kinase inhibitor that was first used to treat Philadelphia chromosome-associated chronic myelogenous leukemia (CML).^2^ This same drug was initially marketed as Gleevec in the US and Glivec in the EU, by the Swiss-American pharmaceutical company Novartis; additional generic brand names now include Celonib, Enliven, Gleevac, Imalek, Imatib, Mesylonib, Mitinab, Plivatinib, Shantinib, Temsan, and Veenat. Before any of these brand names were assigned to the therapeutic, it was published under the identifier ‘STI-571’ in the medical literature.^3^ It can additionally be referenced by the different salt formulations present on the market (imatinib mesylate or imatinib methanesulfonate), or by its chemical structure:

α-(4-methyl-1-piperazinyl)-3’-((4-(3-pyridyl)-2-pyrimidinyl)amino)-p-tolu-p-toluidide Despite their different ontological origins, the preceding examples are all contextually equivalent when referenced with respect to drug-gene or drug-variant interaction annotations, even if there may be subtle distinctions in other, non-therapeutic contexts.

As evidenced from this example, the breadth of nomenclature available for any given therapeutic stems from a variety of contexts relevant to the development and application of that therapeutic. Standards and naming conventions exist at every level of development, from internal pharmaceutical development compound identifiers (ex. AZD-####) and chemical structure names employed in early development pipelines, to fully-realized brand and marketing names with myriad formulations defined by subgroups of additives and delivery mechanisms.^4^ This notion has driven regulatory bodies and programs (such as the United States Adopted Names program) to assign generic names reflecting the underlying active ingredients prior to their marketing. Changes such as these were made in an effort to unify ambiguously named products and protect consumers.^5^ Thus, no matter the stage of development, all assigned names have some tangible link to one another through their relation as a descriptor to the underlying active ingredient(s).

Harmonizing all existing names for any one given therapeutic concept has been a challenging problem for medical informatics in recent decades.^6–8^ While databases and lexical tools exist to address these concerns, many have fallen behind as both the number of therapeutics and the requirements for integration of information resources into medical pipelines continues to evolve and grow.^9–11^ Harmonizing even a single therapeutic requires curated knowledge of all possible identifiers of active ingredients, chemical structures, developmental aliases, and generic or brand names. This also includes all intra-database identifiers such as NCI Thesaurus (NCIt) codes^12^, ChEMBL identifiers^13^, or RxNorm identifiers^9^ (among other resource-based identifiers).

Even when cross-resource identifiers can be utilized properly, there still remains the challenge of maintaining consistent mappings across resources. Stable identifiers are essential for keeping consistent, cross-resource mappings for singular therapeutic concepts. Identifiers lacking a centralized data mapping system may encounter discrepancies among different versions or between users exchanging data. Similarly, how a resource defines individual therapeutics as well as associated aliases can lead to confounding or ambiguous cross-resource mappings. These often occur with therapeutic concepts containing fixed-dose combinations of therapeutics that also exist as singular products. They also occur with products with self-referential components, such as the therapeutic Citalopram. This therapeutic is a racemic mixture of L-citalopram and S-citalopram and runs the risk of producing mappings for its related dextrorotatory therapeutic, escitalopram.

This wealth of possible mappings introduces a great deal of ambiguity and can make communication between different resources challenging. Researchers working with these elements will encounter difficulties when attempting to map therapeutic terms.^4^ Even approximate string matching— itself a computationally-intensive task^14^— is insufficient for capturing linkages between like-therapeutic terms, as variations in formulations may result in entirely different product names. Additional identifiers are needed to ensure proper data harmonization. While these additional identifiers exist, they are often unique to the database housing the information and are largely unstandardized between databases. To bridge this gap, we introduce TheraPy, a Python package and web API that constructs searchable merged concepts for drugs and therapeutic terminologies using publicly available therapeutic resources and thesauri. Merged concepts are constructed from an aggregate set of traits, trade names, and aliases that act as a cross-resource mapping to enable more refined data processing for downstream clinical and research applications. In this report, we outline the methodology behind TheraPy and provide an analysis on normalization rates across different data sources. Further, we examine the challenges of normalization of therapeutic terminologies and provide suggestions on improving data standards to support improving data harmonization.

## RELATED WORK

Ours is not the first methodology to address the problem of normalization of terminology for the biomedical domain. In addition to their exploration of variant, disease, and gene normalization rates, a recent meta-analysis demonstrated a wide disparity in harmonization of therapeutic terminology across six established variant interpretation knowledgebases.^15^ Given the scope and widespread nature of the problem, many other groups have found solutions for normalizing therapeutic concepts within the medical domain. In a recent cross-analysis of precision oncology knowledgebases, Pallarz et al. utilized a synonym set to map filtered drug terms to their matching ChEMBL identifiers^16^. When suitable mappings could not be identified in this manner, manual curation was utilized to resolve conflicts. Expanding on this, others have applied machine learning techniques to identify if two terms are the same concept. Miftahutdinov et al. utilized a modified bioBERT package, to normalize therapeutic vocabulary^17^. In their approach, similarity of entity mentions were optimized with candidate concepts via triplet loss to infer their closest conceptual representation. More similar to our work, Pallarz et al utilized a graph-based approach to normalization^18^. In their work, they created an ontology-based graph for candidate concepts and took advantage of a combined personalized pagerank (PPR) and semantic similarity measure (SSM) to map entity mentions back to their candidates. Our approach expands on existing approaches by taking advantage of established, curated links between concepts across knowledgebases. During the curation process for these knowledgebases, many resources have created crosslinks between singular therapeutic concepts and all their cross-domain semantic representations. By utilizing individual records as nodes, we can then use these crosslinks as edges in our graph-based approach and unify cross-domain vocabulary.

## RESULTS

### Normalization/Grouping Routine

TheraPy utilizes community-driven vocabularies to generate stable concept mappings between identifiers (**Figure 1**). We aggregated concept codes from nine therapeutic ontologies and vocabularies. Terms were extracted from: Wikidata^19^, HemOnc^20^, ChEMBL^13^, the National Cancer Institute Thesaurus^12^ (NCIt), RxNorm^21^, ChemIDplus^22^, Drugs@FDA^23^, DrugBank^24^, and the IUPHAR Guide to Pharmacology.^25^ These sources were chosen due to their high public use as well as the diversity of scope and knowledge contained within each sources. Several of these sources, including RxNorm, Drugbank, NCIt, and HemOnc, directly capture our desired concept space and provide coverage for many molecular and non-molecular therapeutics. Similarly, Drugs@FDA and GuideToPharmacology were utilized to provide additional coverage for active market products and additional therapeutic ligands. Lastly, ChEMBL, ChemIDplus, and Wikidata were included as sources due to their broad coverage of all generalized chemical compounds. Using these sources, we then developed an algorithm to cross-map extracted concept codes and link together records. Normalized identity records are generated in a two-step process:

1. Directed graphs are constructed from source data, where records from each source act as nodes and “has reference to” relationships act as edges between nodes. These relationships are explicit, curated references (xrefs) from one record to another (e.g. the record rxcui:282388 explicitly references drugbank:DB00619) (shown in **Figure 1**).
2. Each set of connected nodes is related as a distinct, unified therapeutic concept and assigned a common identifier. All aliases, trade names, annotations, associations, regulatory approvals and indications are merged under this identifier.

**Figure 1.**
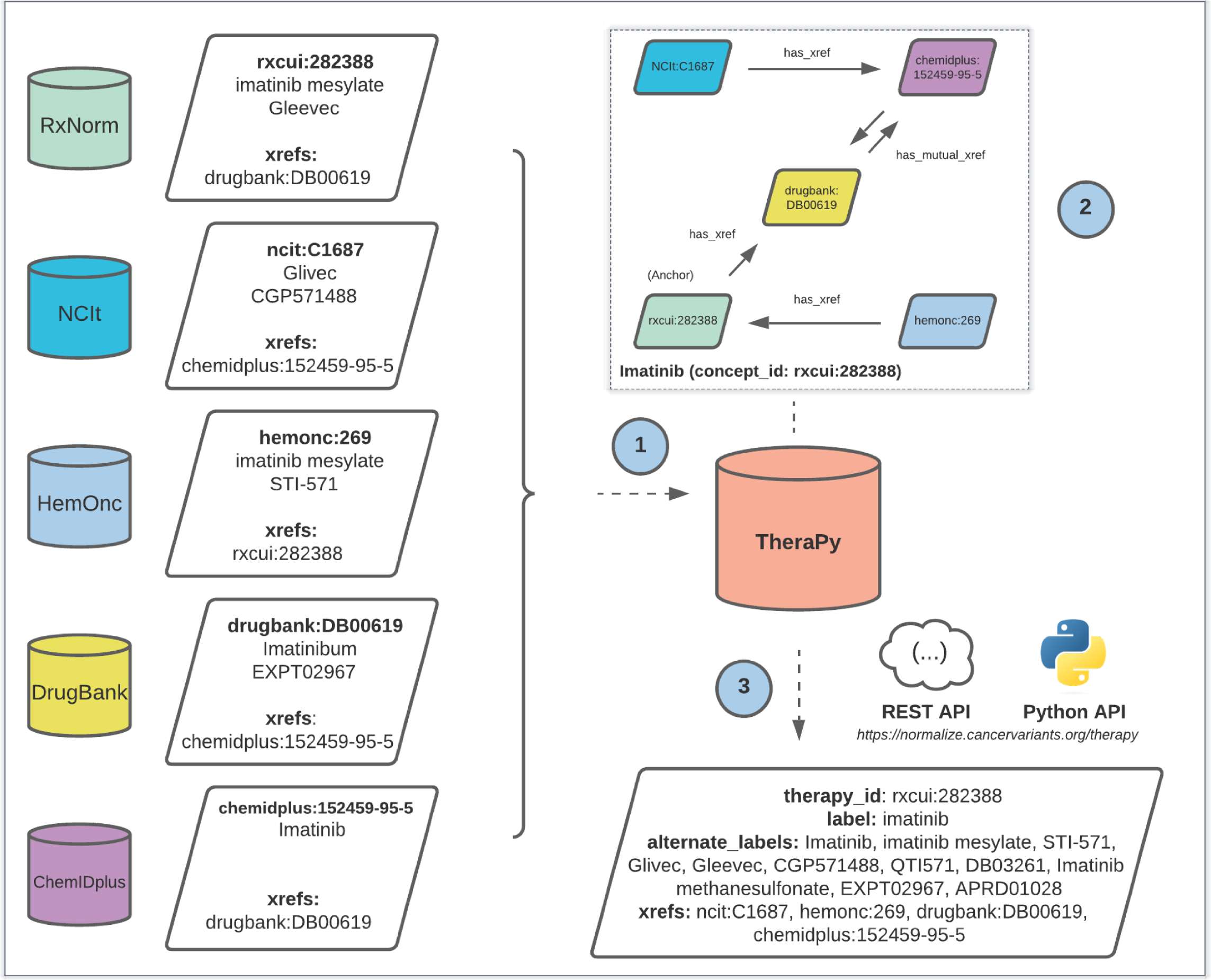
Normalization of therapeutic concepts using TheraPy. Schematic workflow of concept normalization methodology. Shown above is an example workflow for import and normalization of records relating to Imatinib. (1) Therapeutic records are imported from aggregate sources and separated into representative points for all aliases, trade names, explicit cross-references (xrefs), as well as other associations and descriptors. (2) Cross-references are used to draw “has reference to” relationships to similar therapeutics across other sources to create networked groups. The starting node used to create the network is considered the anchor node and becomes the reference identifier for the therapeutic concept. Starting nodes are initialized from an internal source priority hierarchy whereby sources designed for clinical decision making through expert curation were given higher priority than generalized sources. (3) All networked groups are linked under one merged concept record. Raw therapeutic inputs can be normalized to their merged concept record via TheraPy for downstream clinical and research applications. Available from: https://normalize.cancervariants.org/therapy/

Starting nodes were chosen according to an internal source trust ranking, where records with higher priority were used to initialize groups whenever possible. To establish this ranking, sources were ordered according to their perceived therapeutic scope where those designed and annotated primarily for clinical decision making (usually through expert curation) ranked higher than generalized sources. Thusly, the source priority order used for anchor node decision making, from most preferred to least preferred, was: RxNorm, NCIt, HemOnc, Drugbank, Drugs@FDA, IUPHAR Guide to Pharmacology, ChEMBL, ChemIDplus, followed by Wikidata.

### Creation and Access of Normalized Concepts

We ran our normalization routine via TheraPy as described previously in the Methods section. Distinct sets of nodes were assigned a stable merged concept identifier with all associated aliases, trade names, and other therapeutic descriptors associated (**Figure 1**). All merged therapeutic descriptors and cross-references remained accessible via their assigned stable concept identifier.

TheraPy concepts are accessible via HTTP requests to search endpoints, described in our Swagger documentation at https://normalize.cancervariants.org/therapy. In all search types, TheraPy compares the input term with all available concept identifiers, labels, trade names, aliases, cross-references, and associations to find case-insensitive string matches. Matches for the input term are returned in each endpoint and assigned a score corresponding to the type of match identified (Concept ID: 100, Label/Trade Name: 80, Alias/Xref/Association: 60, No Match: 0). Three endpoints are provided to support different kinds of lookups: ’search’, ’normalize’, and ’normalize_unmerged’. The ’search’ endpoint returns all unmerged records with highest available match scores from each individual source given a user input. The ’normalize’ endpoint provides the normalized record for the merged therapeutic concept with the highest available match score given a user query. The ’normalize_unmerged’ endpoint provides the individual source records with the highest available match scores that constitute the normalized concept record given a user query. Further documentation for these endpoints and their associated schemas can be found within our SwaggerUI documentation (https://normalize.cancervariants.org/therapy).

TheraPy is also available as a Python library, installable from the Python Package Interface (PyPI, https://pypi.org/project/therap-py/), and can be run on the command-line. In conjunction with a user-provided DynamoDB database instance (either installed locally or available via Amazon Web Services), data can be loaded by executing the command-line interface module with ‘--update_all’ and ‘--update_merged’ flags. This process will attempt to fetch the latest versions of all source data, extract relevant identifiers, terms, attributes, and cross-references, produce aggregate, normalized terms, and store finished records in the DynamoDB key-value table. Once data loading is complete, TheraPy can be run and queried on a local machine. The project GitHub repository provides additional documentation and basic support for troubleshooting at http://go.osu.edu/TPY.

A total of 16,069 merged groups were created for different therapeutic concepts. These merged groups were assigned identifiers reflective of the anchor node (**Figure 1**) used to create each group: NCIt (6574 groups, 40.9%), RxNorm (4647 groups, 28.9%), GuideToPharmacology (3490 groups, 21.7%), DrugBank (1214 groups, 7.6%), ChEMBL (93 groups, 0.5%), ChemIDplus (51 groups, 0.3%) (**Figure 2a**). Of all merged concepts created, 84.7% of groups contained between 2 and 5 records (**Figure 2b**). The remaining 15.3% of groups contained anywhere from 6 to 86 records. The merged groups with the largest number of combined records are highlighted in **Table 1**.

**Figure 2.**
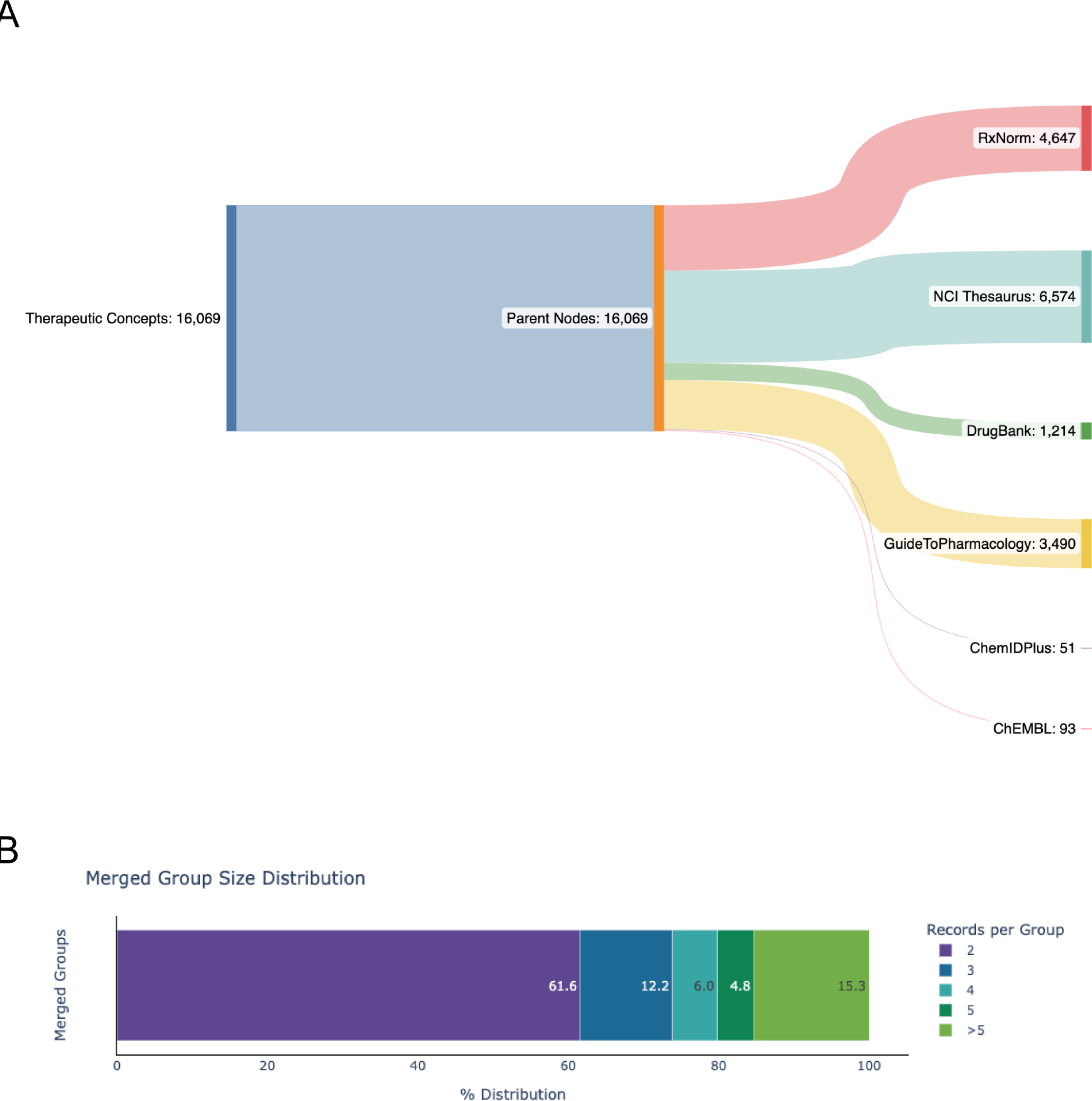
TheraPy normalizes therapeutic records under merged therapeutic concepts. (A) Parent node representation used to assign stable concept identifiers for unique therapeutic concepts. 16,069 unique therapeutic concepts were identified from imported identities. Anchor nodes were selected for each concept from the highest priority available source as defined by our internal priority list. Source priority is represented via verticality (Top: RxNorm, Bottom: ChemIDPlus). **(B)** Distribution of the number of records combined under each unique therapeutic concept.

**Table 1.**
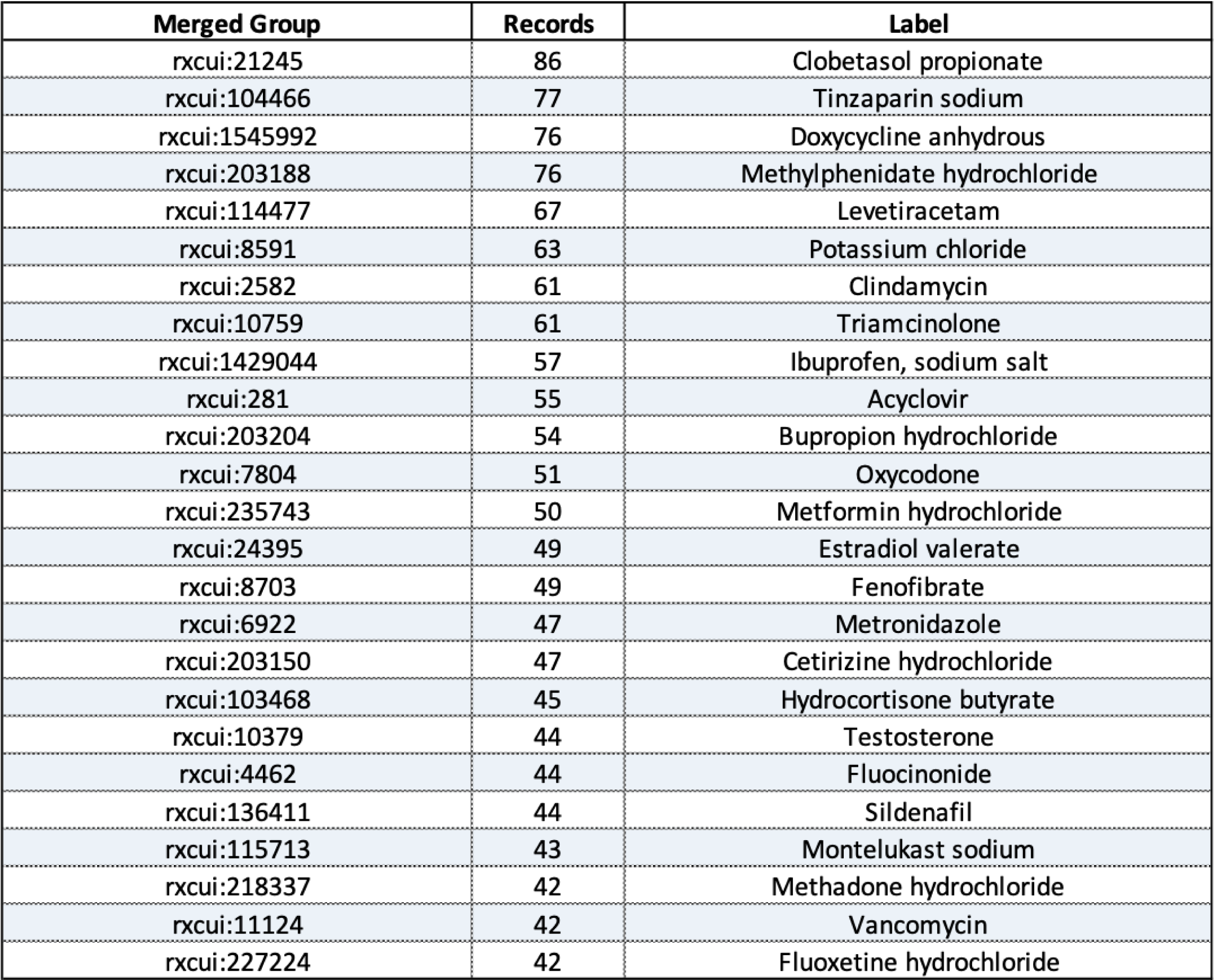
Merged therapeutic concepts with the largest number of combined records within TheraPy. Merged therapeutic concept groups with the most combined records (top 25 shown).

### Analysis of Concept Normalization Rates

To evaluate the ability of TheraPy to successfully harmonize therapeutic terminology across resources, we obtained searchable drug vocabularies from seven different publically available knowledgebases to act as our test set. These knowledgebases are distinct from those used to build TheraPy and are comprised of: Memorial Sloan Kettering (MSK) Precision Oncology Knowledge Base (OncoKB)^26^, Pharmacogenomics Knowledgebase (PharmGKB)^27^, Clinical Interpretation of Variants in Cancer (CIViC)^28^, Cancer Genome Interpreter Cancer Biomarkers Database (CGI)^29^, Molecular Oncology Almanac (MOAlmanac)^30^, Tumor Alterations Relevant for Genomics-Driven Therapy (TARGET)^31^, and the Drug-Gene Interaction Database (DGIdb)^32^. Prior to normalization, therapeutic terminology was compared via string matching to obtain the intersection of common terminology across resources (**Figure 3**). DGIdb was not included in this preliminary analysis due to its nature as an aggregate resource. Our analysis showed a total of 1198 terms unique to a single resource, with 115 and 58 terms being shared across four and five resources respectively.

**Figure 3.**
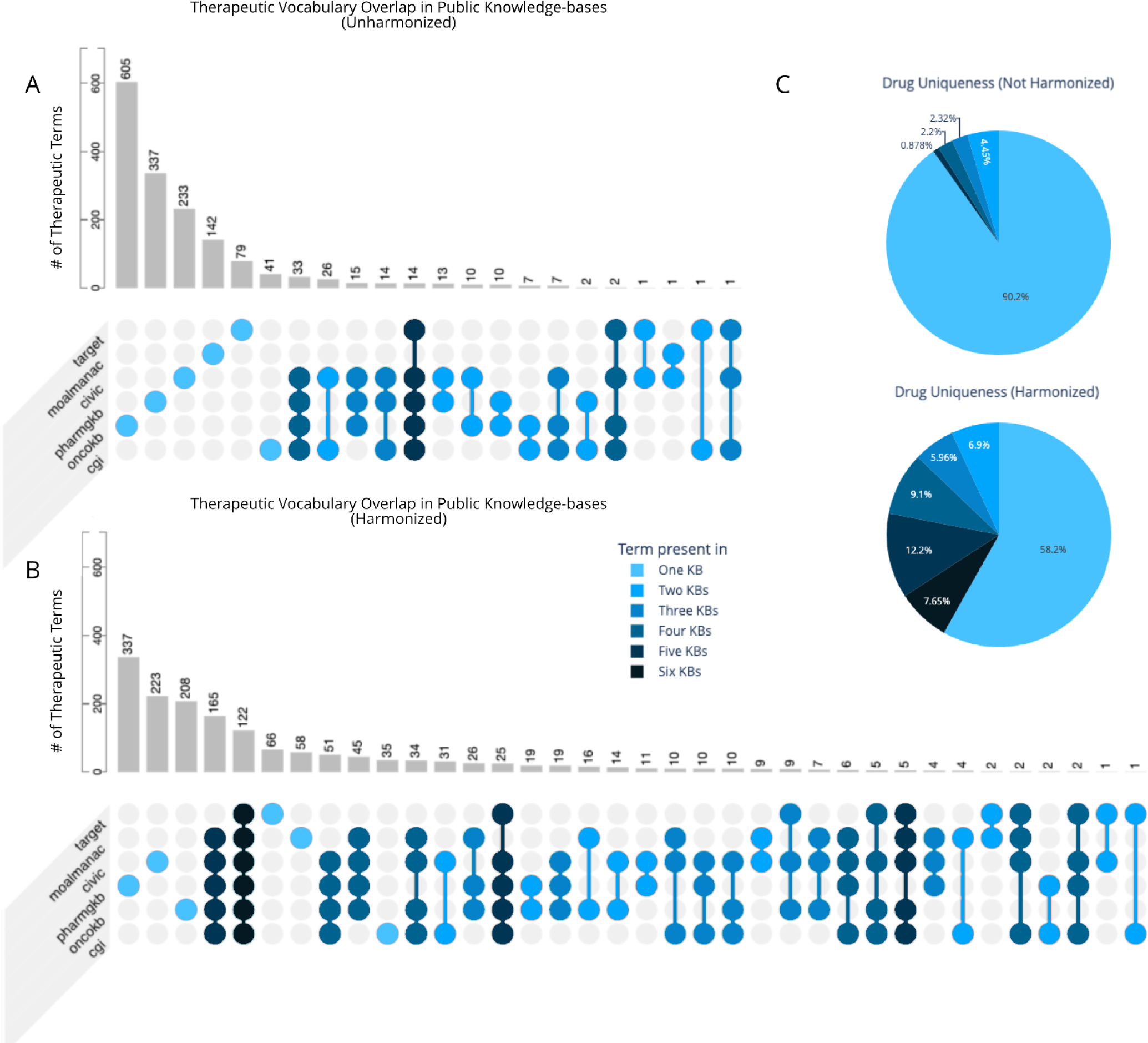
Intersection of therapeutic vocabulary from public knowledge-bases, pre- and post-harmonization using TheraPy. Therapeutic terminology was obtained from six different publicly available drug vocabularies as a test set to evaluate cross-resource therapeutic overlap. (A) Test sets of therapeutic terminology were compared via string matching to quantify the number of exact matches present across resources. The intersections of resources with exact matches are highlighted and colorized by the number of contributing resources. (B) Test sets of therapeutic terminology were harmonized using TheraPy and then compared via concept ID to evaluate the number of matches across resources. Terminologies with exact matches for their concept IDs (irrespective of their original vocabulary term) were quantified. The intersections of resources with matches are highlighted and colorized by the number of contributing resources. (C) Drug uniqueness of therapeutic vocabulary across resources pre- and post-harmonization using TheraPy. Uniqueness is quantified as the number of terms present in various knowledge-bases intersection sizes.

The unique set of terms from each source was then normalized using a local installation of TheraPy (v.0.3.6) and merged concept identifiers were obtained for each term (**Figure 4**). The lack of a merged concept identifier for a unique term was deemed as a failure to normalize. This analysis showed high normalization rates for four of seven sources: PharmGKB (95.6%), CGI (91.2%), OncoKB (86.7%), and CIVIC (85.1%) (**Figure 5a**). The remaining three sources saw lower rates of normalization: MOAlmanac (69.2%), DGIdb (65.4%), and TARGET (36.5%). Examples of terms that failed to normalize are highlighted in **Table 2** and **Table 3**. The anchor nodes for each successfully retrieved merged concept were also recorded. Our analysis of anchor distributions showed RxNorm to be the most frequently-occuring anchor node for drug terms within six out of seven drug sets: OncoKB, PharmGKB, CIVIC, CGI, MOAlmanac, and TARGET (**Figure 5b**). In contrast, ChEMBL was the most frequently occurring anchor node for drug terms obtained from DGIdb.

**Figure 4.**
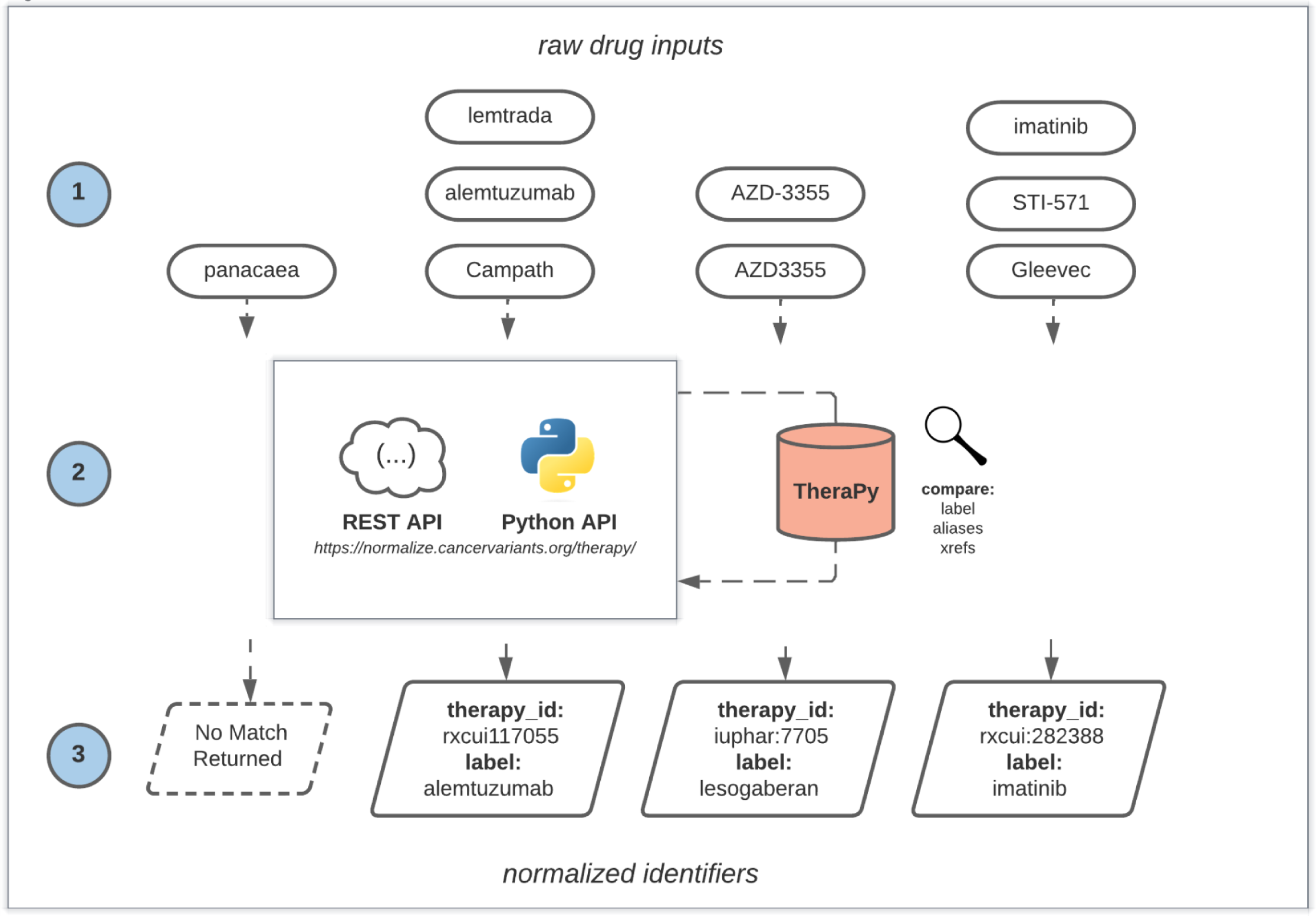
Normalization of therapeutic terminology through TheraPy. Schematic for normalization of therapeutic terms via TheraPy. (1) Raw drug inputs from existing clinical applications are provided to TheraPy as queries. (2) TheraPy compares queried terms (submitted via REST or Python API) against stored aliases, trade names, and other therapeutic descriptors to identify the corresponding merged concepts. (3) TheraPy returns stable merged concepts for downstream usage in clinical applications. If the search term does not exist within the database, no match is returned.

**Figure 5.**
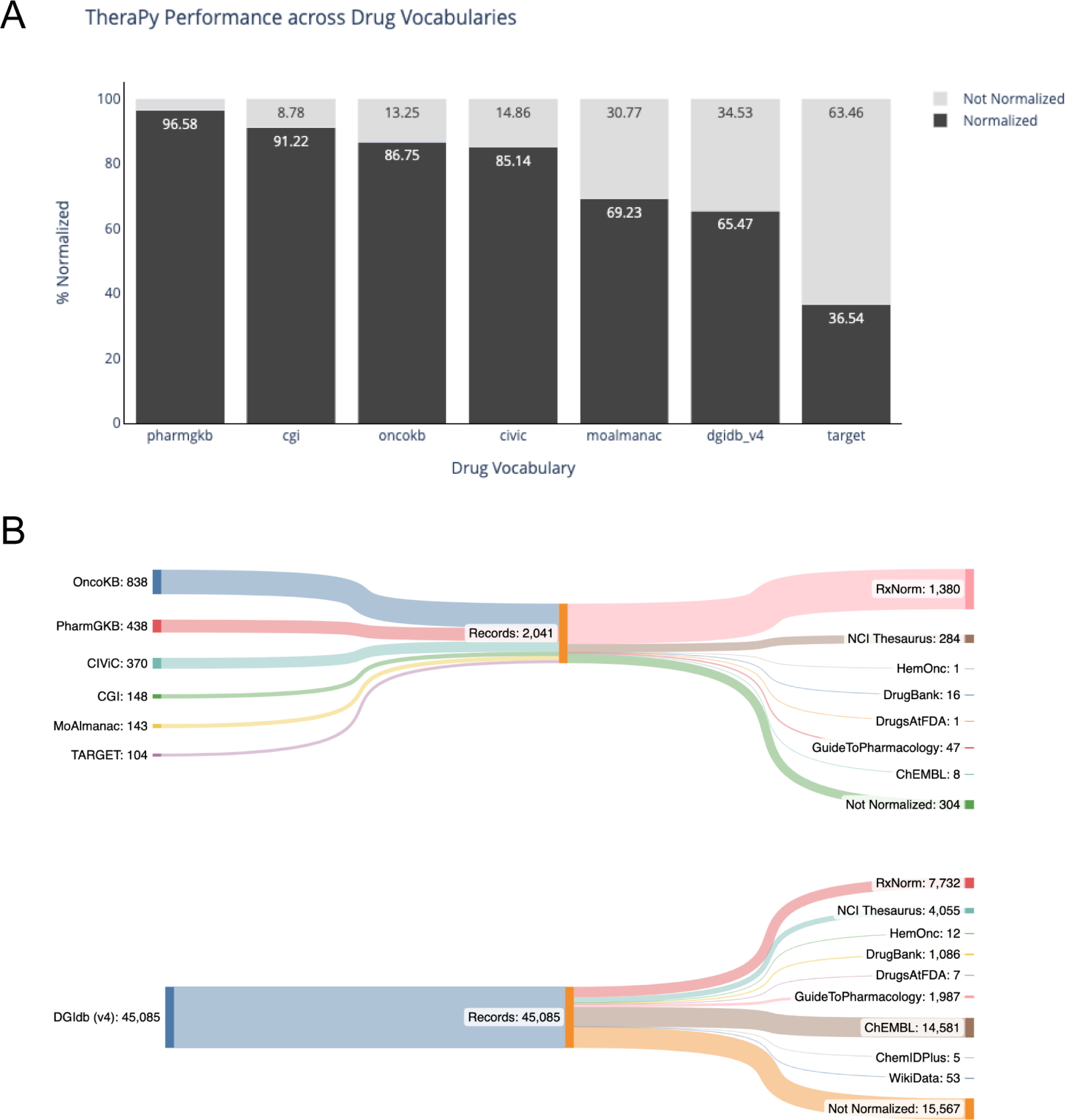
TheraPy normalization performance using publicly available drug vocabularies. (A) Normalization performance for therapeutic terms obtained from seven different publicly available resources. **(B)** Parent node representation for normalized therapeutic terms taken from different resources. Source priority is represented via verticality with records that failed to normalize at the bottom.

**Table 2.**
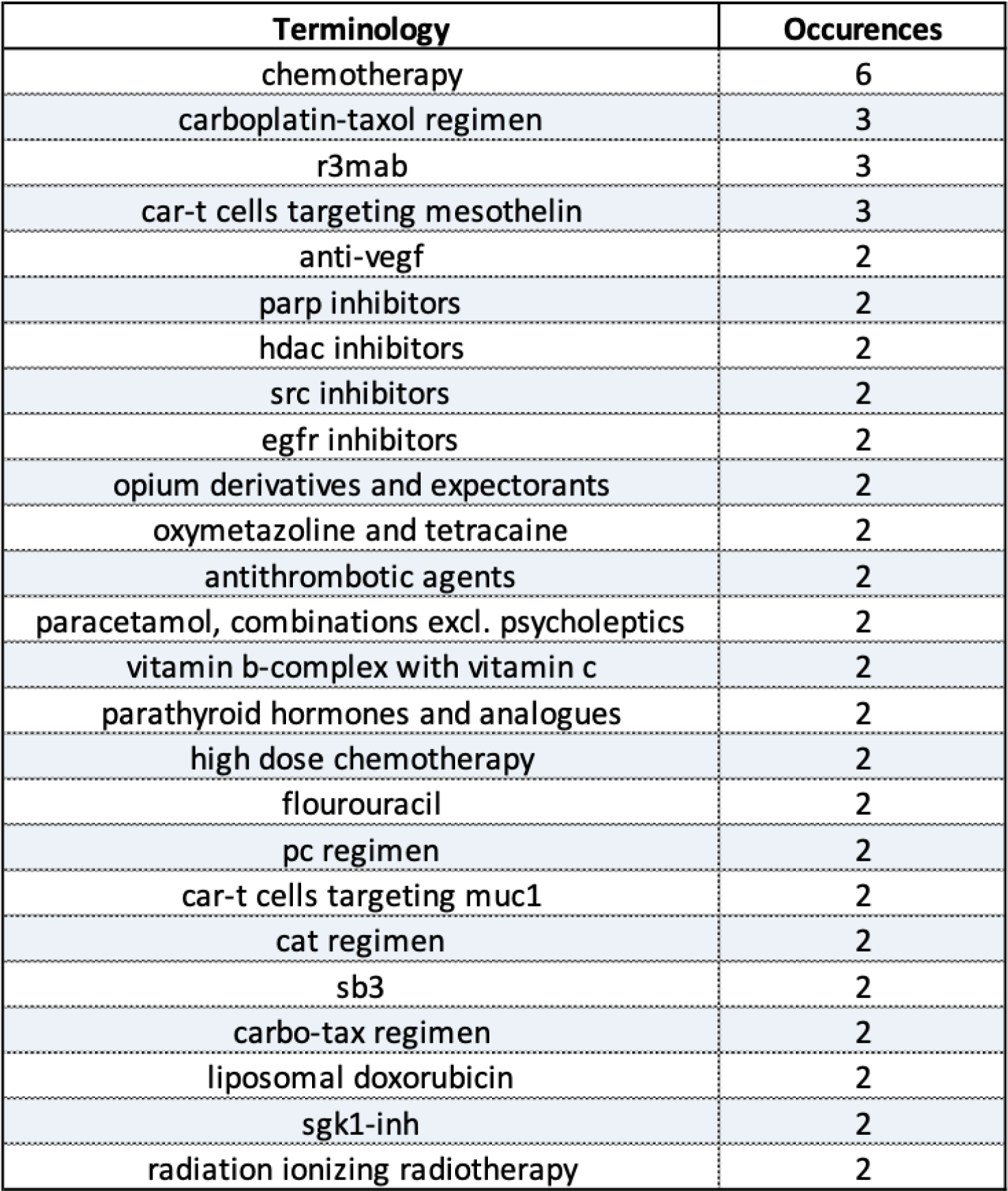
Normalization failure terminology from publicly available drug vocabularies. Most frequently occurring terms (top 25) taken from drug vocabularies that were not able to return matches from TheraPy and thus failed to normalize.

**Table 3.** Additional examples of normalization failure terminology from publicly available drug vocabularies. Select terms taken from drug vocabularies that were not able to return matches from TheraPy and thus failed to normalize. Descriptions of type of drug terminology have been provided next to each term.

| Terminology | Description |
| --- | --- |
| HDAC inhibitors | General Category |
| MEK Inhibitors | General Category |
| beta blocking agents, non-selective | General Category |
| Tyrosine kinase inhibitors | General Category |
| Pyrrolidine derivative 3 | Compound Identifier |
| Carbamate derivative 3 | Compound Identifier |
| Tetra-hydro-isoquinoline derivative 4 | Compound Identifier |
| Benzene sulfonamide derivative 3 | Compound Identifier |
| EVT-103 | Exp. Compound Identifier |
| ADR-851 | Exp. Compound Identifier |
| APN-201 | Exp. Compound Identifier |
| Cysplatyna | Multi-language Label |
| Flourouracil | Misspelled Label |
| Vandetinib | Misspelled Label |
| Radiation Therapy | Therapeutic Description |
| CD19 CAR Gene Transduced T Lymphocytes | Therapeutic Description |
| Anti-PD-L1 CSR T Cells | Therapeutic Description |
| Long-acting erythropoietin conjugate | Therapeutic Description |
| Interferon-alpha lozenge | Therapeutic Description |
| coxsackievirus type a21 | Therapeutic Description |
| 249565746 | Unknown Identifier |

## DISCUSSION

Therapeutic vocabularies from public sources were subjected to directed graph construction to construct stable merged concepts for all descriptors for any given therapeutic concept. Our results showed the construction of 16,069 unique therapeutic concept groups from our import set. We found that 84.7% of all merged concepts created with this methodology contained between 2 and 5 records per group (**Figure 2c**). The remaining 15.3% merged concepts contained >5 records per group with the largest 25 groupings shown in **Table 1**. The size of these larger groupings can likely be attributed to the contributions of one of our sources, Drugs@FDA. The Drugs@FDA resource was added to TheraPy in order to capture more accurate notions of regulatory approval for therapeutic concept groups. In incorporating this resource, TheraPy also links all active Abbreviated New Drug Application (ANDA) and New Drug Application (NDA) records to merged concepts in order to capture all approved products currently on the US market. In doing so, however, this has inflated some therapeutic groups to larger sizes as evidenced by the group ‘rxcui:21245’ containing 84 records (79 of which are ANDA/NDA application records).

Our methodology assigned identifiers reflective of their anchor node based on an established source trust ranking. Sources and vocabularies were ranked according to their therapeutic scope, where sources designed, structured, and annotated primarily for clinical decision making through expert curation were given higher priority than generalized sources with qualification-agnostic curation efforts. This ranking strategy was selected to make terminologies standardized via TheraPy more readily applicable for clinical and genomics applications. It also explains why NCIT, RxNorm, GuideToPharmacology, and DrugBank records were selected as parent nodes for merged concepts more frequently than other imported sources (combined 99%). Interestingly, this also in part explains why ChEMBL was only selected as a parent node for 93 groups (0.5% of groups). Given the scope of ChEMBL as a curated database for all bioactive molecules regardless of clinical usage, this difference may not come as a surprise.

Our analysis of publicly available drug vocabularies showed high rates of normalization for terms obtained from five of seven sources, with TARGET and DGIdb seeing lower rates of normalization for vocabularies (30%, 65.3% respectively). We expect the lower rates of normalization in these two sources to be likely due to more frequent occurrences of general categories (ex. HDAC Inhibitors, MEK Inhibitors), nonspecific identifiers (ex. Pyrrolidine derivative 3, Carbamate derivative 3), misspelled or uncaptured multi-language labels (ex. ‘Vandetinib’, Cysplatyna), or unlisted experimental compound identifiers (ex. EVT-103, ADR-851). Additionally, some terms present within these data sets proved to be therapeutically descriptive but difficult to normalize (ex. CD19 CAR Gene Transduced T Lymphocytes, Anti-PD-L1 CSR T Cells, Long-acting erythropoietin conjugate). While TheraPy does not support fuzzy checks or approximate string matching in its current form, these techniques could be implemented at a later date to handle some of these difficult terminologies. Our normalization results highlight some of the key challenges of therapeutic harmonization. The directed-graph approach behind TheraPy merges descriptors for any therapeutic term listed on a primary node or its cross-referenced edges. While this is sufficient to capture all commonly utilized aliases for a therapeutic, it fails to account for possible variations in spelling that may occur due to language-specific branding or even human error. This is most evident in the failure of TheraPy to normalize the Polish variation of cisplatin, ‘cisplatyna’, or its variant, ‘cysplatyna’. Resolving normalization of terms such as these requires either curation in some form of all possible spelling variations or diligence for correcting human error.

When normalizing therapeutic terms, it is important to carefully consider the nuances of what defines a ‘therapeutic’. Amongst the vocabularies used to create groups and subsequently test TheraPy, we observed many different types of therapeutic categories: natural products, chemical structures, development codes, generic names, brand names, product formulations, and treatment regimens. In our data sets, we observed members of all ontologies co-occurring within the same fields. With all terms carrying a similar weight despite connotations of maturity, it is important to consider these nuances when applying a normalization strategy such as the one we introduced in TheraPy.

We found that our methodology tended to favor normalizing therapeutic concepts to their active ingredient as defined by USAN generic naming standards. Using this strategy, TheraPy was able to reliably capture relationships between the most commonly used therapeutics at the level of generic names, brand names, and even developmental codes or chemical structures in some cases. Conversely, however, it was unable to capture broader therapeutic groupings such as ‘tyrosine kinase inhibitor’ or ‘antibody therapy’. Using our approach, attempts to capture broader descriptors would lead to unintended downstream effects whereby all therapeutics would normalize to their broader therapeutic definition regardless of underlying ingredients (i.e. erlotinib, dasatinib, or gefitinib all normalizing to ‘tyrosine kinase inhibitor’).

As the number of different brands and formulations for available therapeutics continues to grow, so too must the ability for bioinformatic and clinical resources to recognize therapeutic vocabularies. Our results highlight a critical step for harmonizing therapeutic vocabularies in an easily digestible format. By merging available records for any therapeutic concept, we are able to create a therapeutic concept identifier that contains all aliases, trade names, and associations for most commonly used therapeutics. This identifier can be utilized to identify like-therapeutics in different commonly used databases (ex. ChEMBL, RxNorm) and can be incorporated within bioinformatic or clinical workflows to unify therapeutic terms to their underlying active ingredient regardless of maturity stage.

However, more work remains to disambiguate the nuances between therapeutic concept domains. Future effort will require more precise encodings of semantic relations between classes, leveraging recent specifications such as SSSOM for unambiguous, standardized sharing of cross-domain concept mappings. We look forward to these developments, as success in this area will pave the way for applications such as TheraPy to assist inference engines and the development of AI-driven clinical decision support capable of relating disparate therapeutic knowledge resources.

## MATERIALS AND METHODS

### Extraction of Therapeutic Concepts from Resources

Records for drugs, therapeutics, and chemicals were obtained from individual publicly available resources: Terms were extracted from: Wikidata^19^, HemOnc^20^, ChEMBL^13^, the National Cancer Institute Thesaurus^12^, RxNorm^21^, ChemIDplus^22^, Drugs@FDA^23^, DrugBank^24^, and the IUPHAR Guide to Pharmacology.^25^ Further detail on extraction from each individual source can be found below. Records were imported directly as identity records and stored in a locally deployed DynamoDB instance. For each record, aliases, trace names, and database cross-references were extracted and stored as pointers to their original identity.

*ChEMBL* is a “large, open-access drug discovery database that aims to capture Medicinal Chemistry data and knowledge across the pharmaceutical research and development process”^13^, developed by the European Bioinformatics Institute at the European Molecular Biology Laboratory (EMBL-EBI). TheraPy ingests chemical identifiers, labels, aliases, and trade names. Additionally, structured regulatory approval information, including the “max phase” of approval from a survey of regulatory bodies, indicated diseases and phenotypes for use, and withdrawal status if relevant, is acquired. Data is extracted from the latest compressed sqlite distribution (chembl_XX_sqlite.tar.gz) available on the EMBL-EBI FTP server (https://ftp.ebi.ac.uk/pub/databases/chembl/ChEMBLdb/latest/).

*ChemIDplus* was a database storing molecular data about drugs, mechanisms of actions, drug interactions, and drug targets. TheraPy retrieves CAS identifiers, aliases, and cross-references from the most recent ChemIDplus data dump (currentchemid.zip) available on the National Library of Medicine’s FTP server (ftp://ftp.nlm.nih.gov/nlmdata/.chemidlease/).

*DrugBank* is a “web-enabled database containing comprehensive molecular information about drugs, their mechanisms, their interactions and their targets.”^24^ TheraPy extracts drug identifiers, labels, aliases, and cross-references from all the records in the CC0 dataset provided by DrugBank (drugbank vocabulary.csv) in their open data section (https://go.drugbank.com/releases/5-1-10#open-data).

*Drugs@FDA* contains FDA-approved labeling and ingredient information for prescription and over-the-counter drugs, as well as many therapeutic biologics. TheraPy extracts ANDA and NDA marketing application numbers as concept identifiers, as well as trade names, aliases, and some cross-references based on active ingredients. Additionally, the product’s marketing status (discontinued, prescription, over-the-counter, or none) is stored. The Drugs@FDA dataset (drug-drugsfda-0001-of-0001.json.zip) is retrieved from the OpenFDA downloads page (https://open.fda.gov/apis/drug/drugsfda/download/).

*IUPHAR Guide to Pharmacology* is an “open-access, expert-curated database of molecular interactions between ligands and their targets”.^25^ TheraPy makes use of two GtoPdb files: the complete ligand list (ligands.csv) and the ligand ID mapping file (ligand_id_mapping.csv), both available on the GtoPdb downloads page (https://www.guidetopharmacology.org/download.jsp). For each ligand, concept identifiers, labels, aliases, cross-references, and approval data are pulled from the ligand list, and additional cross-references are extracted from the ligand ID mapping file. Approval data reflects whether the drug in question has been permitted for lcinical use by at least one of a selection of regulatory agencies, or whether it has been rmeoved from the market due to safety or other issues.

*HemOnc* is a freely available wiki providing information on therapeutics and protocols in hematology and oncology. A subset of this information is incorporated in the Observational Outcomes Partnership (OMOP) common data model (CDM), enabling its use in “systematic analysis of disparate observational databases… using a library of standard analytic routines and analytic tools”.^20^ TheraPy uses all three files provided as the HemOnc CC-BY subset: YYYY-MM-DD.ccby_concepts.tab, YYYY-MM-DD-ccby_rels.tab, and YYYY-MM-DD-ccby_synonyms.tab. Extracted data includes concept identifiers, labels, trade names, aliases, and cross-references. Additionally, the year of approval by the FDA and a listing of indicated diseases and conditions is acquired.

*NCI Thesaurus* is an initiative by the National Cancer Institute to “integrate molecular and clinical cancer-related information within a unified biomedical informatics framework, with controlled terminology as its foundational layer.”^12^ While its focus is oncology, it has broad coverage of related clinical and scientific knowledge. TheraPy retrieves therapy identifiers, labels, aliases, and cross-references for all classes that descend from “Pharmacologic Substance” (Code C1909), and all classes that have the semantic type “Pharmacologic Substance” but do not have the semantic type “Retired Concept”. Data is pulled using the NCIt Web Ontology Language distribution (Thesaurus_XX.XX.OWL.zip) provided on the NCI FTP server (https://evs.nci.nih.gov/ftp1/NCI_Thesaurus/Thesaurus_23.04d.OWL.zip).

*RxNorm* is a resource provided by the National Library of Medicine to better enable “communicating about clinical drugs and supporting interoperation between drug vocabularies.”^21^ RxNorm is constructed from a number of source terminologies with varying licensing agreements; TheraPy uses information from RxNorm that is provided with a UMLS Source Level Restriction value of 0 or 1. Within those restrictions, TheraPy gathers drug identifiers, aliases, brand and trade names, cross-references, as well as an indication of a drug’s legal prescribable status within the United States. TheraPy collects this data from the latest available RxNorm release (RxNorm_full_MMDDYYYY.zip), provided on the UMLS downloads page (https://www.nlm.nih.gov/research/umls/rxnorm/docs/rxnormfiles.html).

*Wikidata* is an open, collaborative knowledge-base that collects factual claims and descriptions in a Semantic Web model. It is often employed in machine learning and text mining research to provide structured knowledge about a variety of topics. Rather than regular data releases, Wikidata exposes a SPARQL query endpoint for custom requests. TheraPy requests all instances of the class “Medication” and its subclasses, and retrieves IDs, labels, aliases, and available cross-references for each returned class. It saves all data locally in JSON format (as wikidata_YYYY-MM-DD.json) and loads data from there.

### Analysis of Normalization Success Rates

Drug terminology sets were obtained from seven different publically available resources: the Memorial Sloan Kettering (MSK) Precision Oncology Knowledge Base (OncoKB)^26^, Pharmacogenomics Knowledgebase (PharmGKB)^27^, Clinical Interpretation of Variants in Cancer (CIVIC)^28^, Cancer Genome Interpreter Cancer Biomarkers Database (CGI)^29^, Molecular Oncology Almanac (MOAlmanac)^30^, Tumor Alterations Relevant for Genomics-Driven Therapy (TARGET)^31^, and the Drug-Gene Interaction Database (DGIdb)^32^. All drug terms from each source were normalized using a local installation of TheraPy (v.0.3.6). Successful normalization was determined by the retrieval of a merged concept for each term. If a merged concept was not identified, that term was recorded as a failure of normalization.

## Data Availability

All data produced in the present work are contained in the manuscript. Additionally, database and application resources used in the paper are available online for public access at: https://github.com/cancervariants/therapy-normalization

## REFERENCES

1. Quist AJL, Hickman TTT, Amato MG, et al. Analysis of variations in the display of drug names in computerized prescriber-order-entry systems. Am J Health Syst Pharm. 2017;74(7):499–509.

2. Iqbal N, Iqbal N. Imatinib: A breakthrough of targeted therapy in cancer. Chemotherapy Research and Practice, 2014, 1--9. Published online 2014.

3. Imatinib Mesylate (STI-571 Glivec®, Gleevec) Is an Active Agent for Gastrointestinal Stromal Tumours, But Does Not Yield Responses in Other Soft-Tissue Sarcomas That Are Unselected for a Molecular Target: Results from an EORTC Soft Tissue and Bone Sarcoma Group Phase II Study. Excerpta Medica; 2003.

4. Lester CA, Flynn AJ, Marshall VD, Rochowiak S, Rowell B, Bagian JP. Comparing the variability of ingredient, strength, and dose form information from electronic prescriptions with RxNorm drug product descriptions. J Am Med Inform Assoc. 2022;29(9):1471–1479.

5. Karet GB. How Do Drugs Get Named? AMA J Ethics. 2019;21(8):E686–E696.

6. Peters L, Kapusnik-Uner JE, Bodenreider O. Methods for managing variation in clinical drug names. AMIA Annu Symp Proc. 2010;2010:637–641.

7. McCray AT, Srinivasan S, Browne AC. Lexical methods for managing variation in biomedical terminologies. Proc Annu Symp Comput Appl Med Care. Published online 1994:235–239.

8. Eccher C, Ferro A, Pisanelli DM. An Ontology of Therapies. In: Electronic Healthcare. Springer Berlin Heidelberg; 2010:139–146.

9. Nelson SJ, Zeng K, Kilbourne J, Powell T, Moore R. Normalized names for clinical drugs: RxNorm at 6 years. J Am Med Inform Assoc. 2011;18(4):441–448.

10. Dhavle AA, Ward-Charlerie S, Rupp MT, Kilbourne J, Amin VP, Ruiz J. Evaluating the implementation of RxNorm in ambulatory electronic prescriptions. J Am Med Inform Assoc. 2016;23(e1):e99–e107.

11. Khaleel MA, Khan AH, Ghadzi SMS, Alshakhshir S. Curation of an international drug proprietary names dataset. Data Brief. 2022;40:107701.

12. Sioutos N, de Coronado S, Haber MW, Hartel FW, Shaiu WL, Wright LW. NCI Thesaurus: a semantic model integrating cancer-related clinical and molecular information. J Biomed Inform. 2007;40(1):30–43.

13. Gaulton A, Hersey A, Nowotka M, et al. The ChEMBL database in 2017. Nucleic Acids Res. 2017;45(D1):D945–D954.

14. Bringmann K, Künnemann M. Quadratic Conditional Lower Bounds for String Problems and Dynamic Time Warping. In: 2015 IEEE 56th Annual Symposium on Foundations of Computer Science. ; 2015:79-97.

15. Wagner AH, Walsh B, Mayfield G, et al. A harmonized meta-knowledgebase of clinical interpretations of somatic genomic variants in cancer. Nat Genet. 2020;52(4):448–457.

16. Pallarz S, Benary M, Lamping M, et al. Comparative Analysis of Public Knowledge Bases for Precision Oncology. JCO Precision Oncology. 2019;(3):1–8. doi:10.1200/po.18.00371

17. Miftahutdinov Z, Kadurin A, Kudrin R, Tutubalina E. Medical Concept Normalization in Clinical Trials with Drug and Disease Representation Learning. Bioinformatics. Published online July 2, 2021. doi:10.1093/bioinformatics/btab474

18. Lamurias A, Ruas P, Couto FM. PPR-SSM: personalized PageRank and semantic similarity measures for entity linking. BMC Bioinformatics. 2019;20(1):534.

19. Vrandečić D, Krötzsch M. Wikidata: a free collaborative knowledgebase. Commun ACM. 2014;57(10):78–85.

20. Warner JL, Dymshyts D, Reich CG, et al. HemOnc: A new standard vocabulary for chemotherapy regimen representation in the OMOP common data model. J Biomed Inform. 2019;96:103239.

21. Liu S, Ma W, Moore R, Ganesan V, Nelson S. RxNorm: prescription for electronic drug information exchange. IT Prof. 2005;7(5):17–23.

22. Tomasulo P. ChemIDplus-super source for chemical and drug information. Med Ref Serv Q. 2002;21(1):53–59.

23. Center for Drug Evaluation and Research (U.S.). Drugs@FDA. Published 2004. http://www.accessdata.fda.gov/scripts/cder/drugsatfda/

24. Wishart DS, Knox C, Guo AC, et al. DrugBank: a comprehensive resource for in silico drug discovery and exploration. Nucleic Acids Res. 2006;34(Database issue):D668-D672.

25. Harding SD, Armstrong JF, Faccenda E, et al. The IUPHAR/BPS guide to PHARMACOLOGY in 2022: curating pharmacology for COVID-19, malaria and antibacterials. Nucleic Acids Res. 2022;50(D1):D1282–D1294.

26. Chakravarty D, Gao J, Phillips SM, et al. OncoKB: A Precision Oncology Knowledge Base. JCO Precis Oncol. 2017;2017. doi:10.1200/PO.17.00011

27. Whirl-Carrillo M, Huddart R, Gong L, et al. An evidence-based framework for evaluating pharmacogenomics knowledge for personalized medicine. Clin Pharmacol Ther. 2021;110(3):563–572.

28. Griffith M, Spies NC, Krysiak K, et al. CIViC is a community knowledgebase for expert crowdsourcing the clinical interpretation of variants in cancer. Nat Genet. 2017;49(2):170–174.

29. Tamborero D, Rubio-Perez C, Deu-Pons J, et al. Cancer Genome Interpreter annotates the biological and clinical relevance of tumor alterations. Genome Med. 2018;10(1):25.

30. Reardon B, Moore ND, Moore NS, et al. Integrating molecular profiles into clinical frameworks through the Molecular Oncology Almanac to prospectively guide precision oncology. Nat Cancer. 2021;2(10):1102–1112.

31. Van Allen EM, Wagle N, Stojanov P, et al. Whole-exome sequencing and clinical interpretation of formalin-fixed, paraffin-embedded tumor samples to guide precision cancer medicine. Nat Med. 2014;20(6):682–688.

32. Freshour SL, Kiwala S, Cotto KC, et al. Integration of the Drug–Gene Interaction Database (DGIdb 4.0) with open crowdsource efforts. Nucleic Acids Res. 2020;49(D1):D1144–D1151.

